# Temozolomide sensitivity of malignant glioma cell lines – a systematic review assessing consistencies between *in vitro* studies

**DOI:** 10.1101/2021.06.29.21259733

**Authors:** Michael TC Poon, Morgan Bruce, Joanne E Simpson, Cathal J Hannan, Paul M Brennan

## Abstract

Malignant glioma cell line models are integral to pre-clinical testing of novel potential therapies. Accurate prediction of likely efficacy in the clinic requires that these models are reliable and consistent. We assessed this by examining the reporting of experimental conditions and sensitivity to temozolomide in glioma cells lines.

We searched Medline and Embase (Jan 1994-Jan 2021) for studies that evaluated the effect of temozolomide monotherapy on cell viability of at least one malignant glioma cell line. Studies using a drug-resistant cell line or a modified preparation of temozolomide were excluded. Key data items included type of cell lines, temozolomide exposure duration, and cell viability measure (IC_50_).

We included 212 eligible studies from 2,789 non-duplicate records that reported 248 distinct cell lines. The commonest cell line was U87 (60.4%). Only 10.4% studies used a patient-derived cell line. The proportion of studies not reporting each experimental condition ranged from 8.0-27.4%, including base medium (8.0%), serum supplementation (9.9%) and number of replicates (27.4%). In studies reporting IC_50_ the median value for U87 cell line at 24 hours, 48 hours and 72 hours was 123.9μM (IQR 75.3-277.7μM), 223.1μM (IQR 92.0-590.1μM) and 230.0μM (IQR 34.1-650.0μM), respectively (Figure 2A). The median IC_50_ at 72 hours for patient-derived cell lines was 220μM (IQR 81.1-800.0μM).

Temozolomide sensitivity reported in comparable studies was not consistent between and within individual malignant glioma cell lines. Drug discovery science performed on these models cannot reliably inform clinical translation. A consensus model of reporting can maximise reproducibility and consistency among *in vitro* studies.

**Key points:**

- There is a wide variety of study designs for malignant glioma cell line studies.
- Reporting of experimental designs of cell line studies was suboptimal.
- Temozolomide sensitivity was inconsistent between and within individual cell lines.

**Importance of the study:**

- There is a wide variety of experimental designs for malignant glioma cell line studies but the reporting of these is suboptimal.
- Temozolomide sensitivity reported in comparable studies was not consistent between and within individual malignant glioma cell lines.
- While there will be variations of opinion on what the optimal design is, a consensus model of a reporting structure is the only rational way to maximise the yield from *in vitro* studies to find novel therapies for our patients.

## Introduction

Malignant glioma is the most common primary cancer of the central nervous system.^1^ Treatment options vary between types of malignant glioma, but prognosis is poor.^2^ Classification depends on histological and molecular features.^3^ Glioblastoma, accounting for 60% of all malignant gliomas, carries the worst outcome with median survival of 6-8 months and 3% 5-year survival.^2,4,5^ Standard care has not changed in 15 years and involves surgical resection followed by a combination of radiotherapy and chemotherapy with the alkylating agent temozolomide.^6^ No human clinical trials to date have demonstrated superior treatment effect with a novel therapeutic agent,^7^ so there is an urgent need to examine the processes of drug discovery.

Cancer drug discovery utilises pre-clinical models to predict the likely effect of a novel compound on cancer tissue. *In vitro* cell cultures are readily available from repositories^8^ and in studies of malignant glioma, temozolomide is frequently used as the comparative therapeutic agent because of its role in standard care. The robustness of this strategy depends on the reproducibility of temozolomide sensitivity *in vitro*, which may be affected by variations in experimental conditions.^9,10^ This is critical to experimental design and to data interpretation, so requires adequate reporting.

Transparency in clinical research has received increasing attention in the past two decades. There has been an expansion of reporting guidelines for clinical studies facilitated by the Enhancing the Quality and Transparency Of health Research (EQUATOR) network.^11^ Most guidelines relate to clinical research, but the Animal Research: Reporting of In Vivo Experiments (ARRIVE) guidelines are relevant to pre-clinical animal models.^12^ However, there are currently no standardised reporting guidelines for cell culture studies. Evaluation of the experimental conditions and drug sensitivity of cell lines in published reports may provide an insight into the reproducibility and consistency in these models. The aim of this review was to quantify the variability and reporting of experimental conditions in *in vitro* studies using malignant glioma cell lines and to assess variations in temozolomide sensitivity.

## Methods

There is no suitable repository for systematic review of *in vitro* studies in which our protocol could be registered. We followed the Preferred Reporting Items for Systematic Reviews and Meta-Analyses (PRISMA) guidelines.

### Eligibility criteria

We included studies published after 1993 that evaluated the effect of temozolomide monotherapy on cell viability of at least one malignant glioma cell line. The evaluation of temozolomide could be a control experiment within a study. There was no limit on the type of cell viability measure. We excluded studies that utilised a drug-resistant cell line, a modified preparation of temozolomide, or an animal model. Studies using cell line-based xenotransplant animal models or reporting outcome measures other than cell viability were excluded.

### Information sources

We searched Ovid Medline and Embase between January 1994 and January 2021 using a combination of search terms relating to gliomas, cell lines, temozolomide, and cell death. The full search strategies are available in Supplementary Material. Our search date was on 28^th^ January 2021. There was no hand search or search in the grey literature.

### Study selection & data extraction

The online tool Covidence was our platform for conducting the primary screening and data extraction in this review. We applied the built-in deduplication function on the platform to records retrieved from our database search. One reviewer (MB) screened titles and abstracts of all records, of which 10% excluded records were reviewed by a second reviewer (MTCP). We used the same review approach for full-text eligibility assessment. One reviewer (MB) extracted data from all eligible articles and a second reviewer (MTCP) independently extract data from 40% of these studies. We resolved disagreements by discussion between the two reviewers or by seeking resolution from a third reviewer (PMB).

### Data items

We collected data on study characteristics, experimental setup and cell viability measures. Study characteristics included year of publication, country of primary affiliation, the primary aim of study, and types of cell lines used. ‘Primary aim of study’ had three categories: “therapeutic evaluation” referred to studies comparing the effects on cell lines of another therapeutic agent alone against temozolomide; “pathway modification” referred to studies that altered cellular pathways with a molecularly targeted compound and assessed the effect of temozolomide on the cell lines; “gene or protein measurement” referred to studies that measured levels of molecular markers in response to temozolomide. We categorised cell lines into human cell lines, murine cell lines, and cancer stem cell-like patient-derived cell lines. Patient-derived cell lines were developed from patients with malignant glioma at the investigators’ institute. Experimental setup data included temozolomide concentration, temozolomide exposure duration, base medium, addition of serum, cell density, cell passage number, use of hypoxic environment, and temperature and percentage of carbon dioxide in the ambient incubator environment.

Cell viability was our outcome of interest. Data on cell viability included the assay, quantification technique used and cell viability measurement. Cell viability assays included 3-(4,5-dimethylthiazolyl-2)-2,5-diphenyl-2H-tetrazolium bromide (MTT), sulphondamine B (SRB), annexin V, propium iodide (PI), trypan blue, bromodeoxyuridine (BrdU) and luciferase-based techniques. Quantification techniques included flow cytometry, colorimetric or fluorometric plate reader, microscopy with manual counting, and microscopy with automated counting. Where reported, we collected drug sensitivity measures on half-maximal inhibitory concentration (IC_50_), half-maximal effective concentration (EC_50_), dose reduces initial population to 10% (LD_10_), dose reduces survival number of cells to 37% (D_37_), approximate concentration of drug tolerated without lethality (DT), and median-effect dose (Dm). To assess the internal validity, we assessed whether a study reported the number of replicate plates and the number of replicates per plate.

### Risk of bias

There was no suitable risk of bias tool for the type of studies included in this review. The completeness of reporting of the data items described above was instead used to indicate the transparency of the studies to permit summarisation of comparable studies.

### Data synthesis

We used descriptive statistics to summarise the data items. Our reporting of temozolomide sensitivity measures was stratified for the two most common cell lines (U87, U251) and patient-derived cell lines. We divided studies into groups that had the same cell lines exposed to temozolomide for the same duration, in a non-hypoxic environment, and reported the same drug sensitivity measures. We tested the homogeneity of variances in temozolomide sensitivity across cell lines using Levene’s and Fligner-Killeen tests. There was no plan for meta-analysis because studies were anticipated to be heterogeneous and drug sensitivity measures without a measure of variance are not amenable to meta-analysis. These reasons also precluded the assessment of reporting bias. We performed all analyses in R version 4.1.0 using ‘*tidyverse*’ (v1.3.1), ‘*gtsummary*’ (v1.4.1), and ‘*car*’ (v.3.0-10) packages.

## Results

### Study selection and characteristics

Our search retrieved 3,586 records. After removing 797 duplicates, 2,789 records underwent title and abstract screening, of which 1,532 studies had full-text eligibility assessment. We included 212 eligible studies^s1-213^ using malignant glioma cell lines that reported cell viability measures associated with temozolomide (Figure 1). Of the 1,320 studies excluded at full eligibility assessment, the primary reason for exclusion for 717 studies was no reporting of cell viability measures.

**Figure 1.**
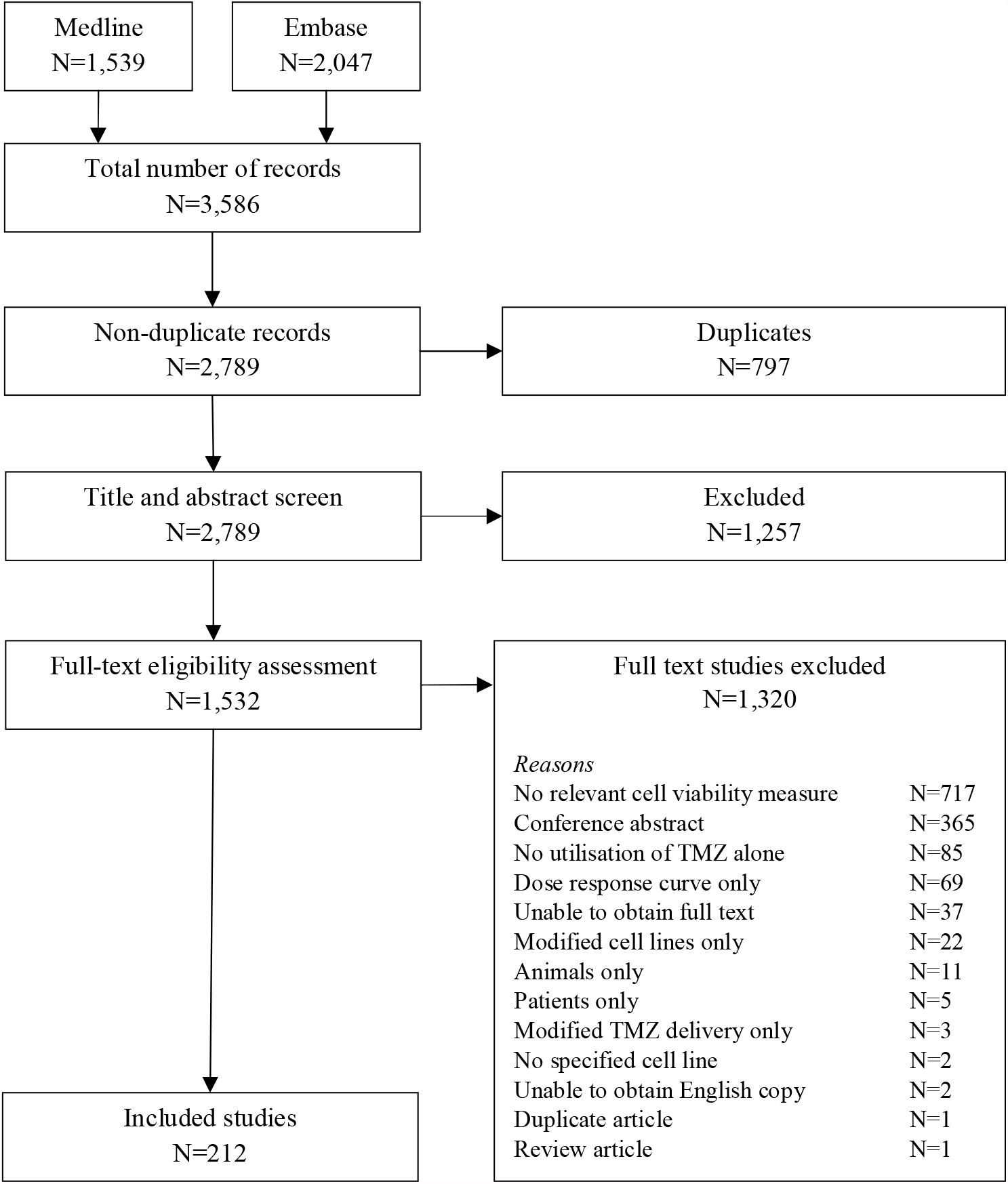
Flowchart describing study selection for inclusion

Of all the included studies, 140 (66.0%) were published in or after 2015. Countries accounting for over 10% of the included studies were China (35.4%) and the United States (17.9%). The proportion of studies published in or after 2015 from China and from the United States was 78.9 % and 57.9%, respectively. Most studies (85.4%) evaluated therapeutic agents against temozolomide in their cell lines as the primary aim. There were 19 (9.0%) studies measuring gene or protein levels in their cell lines in response to temozolomide and 12 (5.6%) studies assessed the cell line response to temozolomide after modification of a cellular pathway. A human glioma cell line was at least one of the cell lines used in 193 (91.0%) studies. Twenty-three (10.8%) studies used at least one murine cell line and 22 (10.4%) studies used at least one patient-derived cell line (Table 1). Most studies (77.4%) used two or more cell lines in their studies.

**Table 1.** Characteristics of malignant glioma cell line reported in 212 included studies

| Characteristics | Year of publication |  |  |  |
| --- | --- | --- | --- | --- |
|  | Overall<br>N = 212 | Before 2015<br>N = 72 | 2015-2017<br>N = 63 | 2018 or after<br>N = 77 |
| Type of cell line |  |  |  |  |
| Human glioma cell line(s) | 193 (91.1%) | 65 (90.3%) | 57 (90.5%) | 71 (92.2%) |
| Murine glioma cell line(s) | 23 (10.8%) | 5 (6.9%) | 6 (9.5%) | 12 (15.4%) |
| Patient-derived cell line(s) | 22 (10.4%) | 9 (12.5%) | 7 (11.1%) | 6 (7.7%) |
| Number of cell lines used |  |  |  |  |
| One | 48 (22.6%) | 14 (19.4%) | 15 (23.8%) | 19 (24.7%) |
| Two | 74 (34.9%) | 19 (26.4%) | 26 (41.3%) | 29 (37.7%) |
| Three or more | 90 (42.5%) | 39 (54.2%) | 22 (34.9%) | 29 (37.7%) |
| Ten commonest cell lines |  |  |  |  |
| U87 | 129 (60.4%) | 47 (65.3%) | 37 (58.7%) | 44 (57.1%) |
| U251 | 87 (41.0%) | 24 (33.3%) | 31 (49.2%) | 33 (41.6%) |
| T98G | 56 (26.4%) | 25 (34.7%) | 11 (17.5%) | 20 (26.0%) |
| A172 | 29 (13.7%) | 8 (11.1%) | 11 (17.5%) | 10 (13.0%) |
| U373 | 27 (12.7%) | 15 (20.8%) | 6 (9.5%) | 6 (7.8%) |
| U138 | 26 (12.3%) | 15 (20.8%) | 6 (9.5%) | 5 (6.5%) |
| LN229 | 22 (10.4%) | 7 (9.7%) | 4 (6.3%) | 11 (14.3%) |
| C6 | 20 (9.4%) | 5 (6.9%) | 6 (9.5%) | 9 (11.7%) |
| LN18 | 13 (6.6%) | 6 (8.3%) | 2 (3.2%) | 5 (6.5%) |
| U118 | 14 (6.6%) | 5 (6.9%) | 5 (7.9%) | 4 (5.2%) |

### Cell lines & experimental setup

Of 750 cell lines reported in the 212 included studies, there were 248 distinct cell lines. The three most used cell lines were U87 (60.4%), U251 (41.0%), and T98G (26.4%). Their use did not appear to change over time (Table 1). Patient-derived cell lines were used in 7.8% studies published in 2018 or after compared with over 12.5% in studies published before 2015. There were 130 distinct patient-derived cell lines reported in 22 studies.

Dulbecco’s Modified Eagle Medium (DMEM) was the commonest base medium, used in 142 (66.5%) studies (Table 2). There were 23 (10.8%) studies that used a customised medium and 17 (8.0%) studies did not report the type of base medium used. Reporting of carbon dioxide concentration and temperature was missing in 54 (25.5%) and 52 (24.5%) studies, respectively. Five (2.3%) studies utilised a hypoxic environment, but 54 (25.5%) studies did not report the oxygen specification. Cell density was frequently reported in 154 (72.6%) studies. Cell passage number was reported in only 16 (7.5%) studies.

**Table 2.**
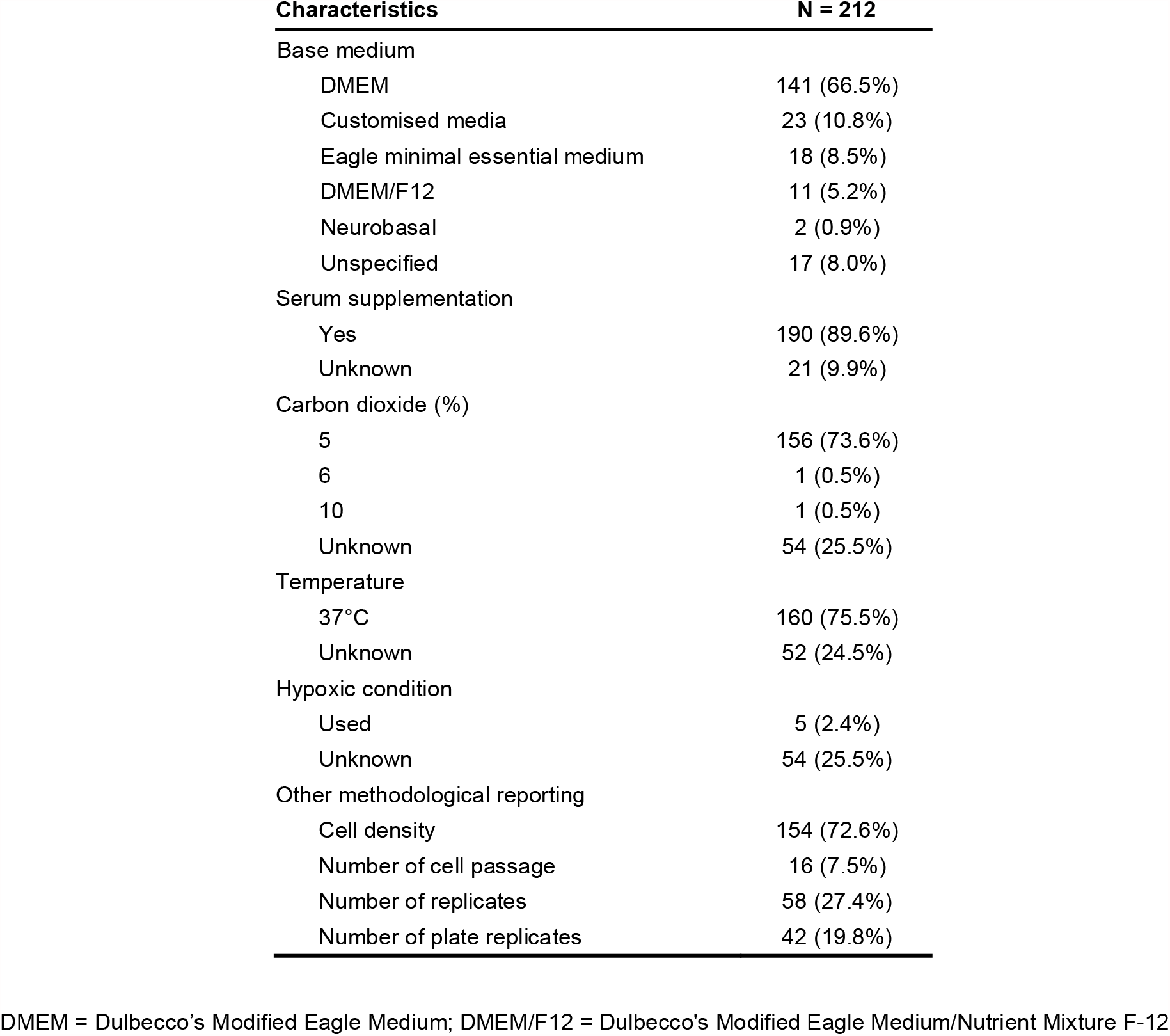
Experimental design of 212 included studies

### Specification of temozolomide use

The range of temozolomide concentrations used in determining drug sensitivity was not determinable from 30 (14.2%) studies (Table 3). The median lower limit of temozolomide concentration was 0.1μM (range 0-6,250μM, interquartile range [IQR] 0-10μM) and the median upper limit was 800μM (range 30-400,000μM, IQR 250-1,266μM). A single temozolomide exposure time was not reported in 47 (22.2%) studies, of which 38 studies instead reported a range of exposure times. The three most common temozolomide exposure times were 72 hours (29.1%), 48 hours (16.7%), and 24 hours (11.8%).

**Table 3.** Temozolomide specification and methods for measuring cell viability

| Characteristics | N = 212 |
| --- | --- |
| Temozolomide concentration ($\mu\text{M}$) | |
| Lower range (IQR)<sup>†</sup> | 0 (0-10) |
| Upper range (IQR)<sup>†</sup> | 800 (250.0-1266) |
| Unspecified | 30 (14.2%) |
| Temozolomide exposure time |  |
| <24 hours | 8 (3.8%) |
| 24 hours | 24 (11.3%) |
| 48 hours | 34 (16.0%) |
| 72 hours | 59 (27.8%) |
| 96 hours | 15 (7.1%) |
| >96 hours | 25 (11.8%) |
| Range of time | 38 (17.9%) |
| Unspecified | 9 (4.2%) |
| Cell viability assay |  |
| MTT | 95 (44.8%) |
| CCK-8 | 35 (16.5%) |
| MTS | 11 (5.2%) |
| Trypan blue | 11 (5.2%) |
| Other | 55 (25.9%) |
| Unspecified | 5 (2.4%) |
| Method for cell viability quantification |  |
| Plate reader | 124 (58.5%) |
| Flow cytometry | 13 (6.1%) |
| Microscopy and manual counting | 11 (5.2%) |
| Microscopy and automated counting | 3 (1.4%) |
| Technique in a reference/manual | 16 (7.5 %) |
| Unspecified | 45 (21.2%) |

<sup>†</sup>Range of temozolomide used to assess drug sensitivity was reported in 183 studies. IQR = interquartile range; MTT = 3-(4,5-dimethylthiazolyl-2)-2,5-diphenyl-2H-tetrazolium bromide; CCK-8 = Cell Counting Kit-8; MTS = MTT with phenazine methosulfate

### Cell viability methods and drug sensitivity measures

MTT was the most common primary cell viability assay, which was used in 95 (44.8%) studies (Table 3). Thirty-five (16.5%) studies used the Cell Counting Kit-8, another colorimetric assay. Five (2.4%) studies did not specify their cell viability assays. A plate reader was used in 124 (58.5%) studies and 45 (21.2%) studies did not specify their quantification techniques for measuring cell viability. Among our pre-specified drug sensitivity measures, 182 (85.8%) studies reported IC_50_ of temozolomide for their cell lines. The other drug sensitivity measures each had fewer than 10 studies reporting them.

### Temozolomide sensitivity

We examined studies together that used the same temozolomide exposure time and reported IC_50_ of temozolomide for their cell lines (Figure 2). Descriptive statistics here relate to cell lines at each exposure duration that had ≥10 measurements of temozolomide IC_50_. The median IC_50_ of temozolomide for U87 cell line at 24 hours, 48 hours and 72 hours was 123.9μM (IQR 75.3-277.7μM), 223.1μM (IQR 92.0-590.1μM) and 230.0μM (IQR 34.1-650.0μM), respectively (Figure 2A). In U251 cell line, the median temozolomide IC_50_ at 48 hours and 72 hours was 240.0μM (IQR 34.0-338.5μM) and 176.50μM (IQR 30.0-470.0μM), respectively. The median IC_50_ of temozolomide at 72 hours for T98G and patient-derived cell lines was 438.3μM (IQR 232.4-649.5μM) and 220μM (IQR 81.1-800.0μM), respectively.

**Figure 2.**
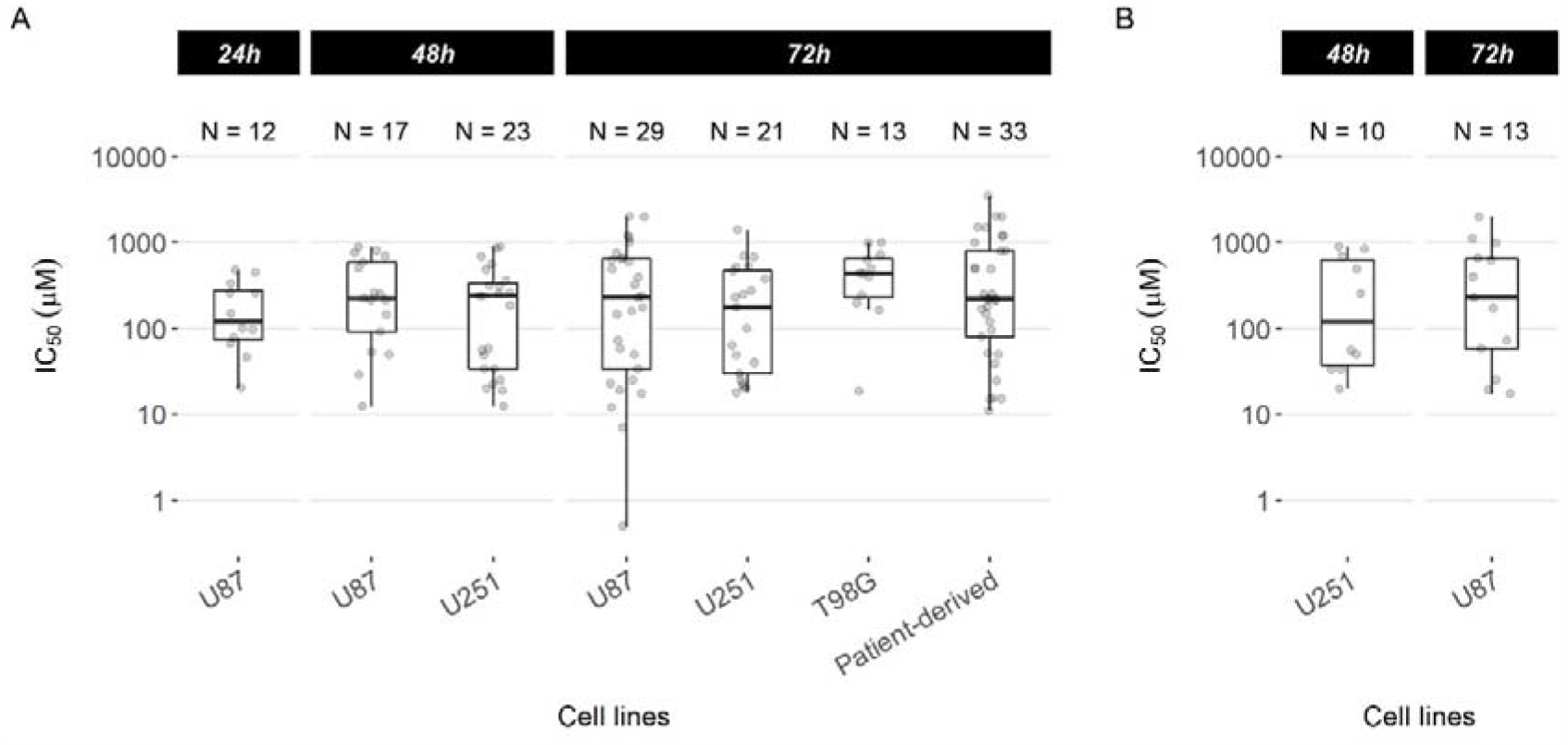
Temozolomide sensitivity measured by half-maximal inhibitory concentration (IC_50_) in different cell lines stratified by temozolomide exposure time. (A) IC_50_ summarised for cell lines that had at least 10 measurements reported in the included studies at the specified exposure time. Tests for homogeneity of variances showed no evidence of a difference between cell lines at 72 hours (Levene ‘s test: p=0.249; Fligner-Killeen test: p=0.389). References for these studies are: U87 at 24hr,^s9 s21 s26 s56 s57 s60 s83 s91 s105 s120 s146 s209^ U87 at 48hr,^s31 s58 s67 s88 s96 s109 s112 s114 s121 s156 s160 s163 s189 s197 s205 s209 s211^ U251 at 48hr,^s17 s44 s29 s31 s58 s67 s88 s96 s99 s109 s112 s114 s116 s149 s152 s156 s175 s184 s186 s187 s189 s197 s211^ U87 at 72hr,^s7 s13 s14 s48 s50 s63 s94 s97 s98 s104 s106 s128 s130 s134 s135 s137 s138 s145 s154 s158 s166 s169 s174 s195 s196 s198 s200 s203 s212^ U251 at 72hr,^s2 s30 s37 s46 s59 s97 s125 s131 s134 s135 s145 s154 s166 s167 s174 s181 s185 s196 s200 s203 s212^ T98G at 72hr,^s2 s28 s46 s48 s63 s94 s145 s166 s167 s192 s196 s200^ and patient-derived cell lines at 72hr.^s14 s122 s123 s127^ (B) Further restricting to studies that did not use a hypoxic environment and quantified cell viability using MTT. Exposure time in hours is shown on the top panel. IC_50_ = half-maximal inhibitory concentration. References for these studies are: U251 at 48hr ^s31 s44 s58 s96 s149 s152 s156 s175 s187 s189^ and U87 at 72 hours.^s7 s13 s98 s106 s128 s130 s134 s137 s145 s166 s195 s200 s212^

The asymmetrical distribution of the IC_50_ values demonstrated in Figure 2A may be caused by the differing experimental conditions. To examine for the source(s) of this variation in IC_50_, we then included studies that used a normoxic culture environment (excluding hypoxic culture condition) and that used MTT as the cell viability assay, the most common composite conditions. The IC_50_ was 155.1μM (IQR 37.8-640.5μM) for U251 cell line at 48 hours and 230.0μM (IQR 58.0-650.0μM) for U87 cell line at 72 hours (Figure 2B), compared to 240.0μM and 230.0μM in the previous analysis of U251 and U87, respectively, at the same time points. The interquartile ranges of the IC_50_ from more homogeneous studies and from unrestricted studies were similar.

Tests for homogeneity did not demonstrate evidence of a difference in IC_50_ variances across cell lines exposed to temozolomide for 72hrs (Levene’s test: p=0.237; Fligner-Killeen test: p=0.346).

## Discussion

This systematic review identified 213 studies using cell culture models of malignant gliomas reporting drug sensitivity to temozolomide. Over 700 studies were excluded because they did not report cell viability measures. There were a wide variety of cell lines, and we did not observe an increase over time in patient-derived cell lines despite the implications of the cancer stem cell hypothesis for glioma biology. The reporting of experimental conditions and temozolomide specifications were variable, which made interpretation difficult. Temozolomide sensitivity was not consistent within each cell line. Furthermore, the range of concentrations used across the selected publications is not physiologically or clinically relevant to glioblastoma treatment (approximately 30-50uM in tumour and plasma^13^). This has implications for conduct and interpretation of drug discovery.

Sampling variation of temozolomide sensitivity in the same cell lines between studies should produce a symmetrical distribution of IC_50_ around the median. The asymmetrical distributions of IC_50_ in each cell line (Figure 2A) suggested additional contributors to inconsistency other than sampling variations. While further matching of experimental conditions (Figure 2B) reduced the asymmetry, the residual asymmetry indicated that inconsistency resulted from differences in unmeasured experimental conditions and in cell behaviours in each cell line. Issues with cultured malignant glioma cell lines are not new. Serum supplementation in classic cell lines has been shown to induce astrocytic differentiation resulting in transcriptional and epigenomic changes that do not reflect the human disease.^14,15^ There are further problems with misidentification and cross-contamination.^16,17^ These limitations can contribute to an explanation of our findings of cell lines having inconsistent temozolomide sensitivity. Drug discovery science performed on these incomparable and nonrepresentative malignant glioma models cannot reliably inform clinical translation.

Although there was no apparent increase in its use, about a quarter of the included studies used patient-derived glioma cell lines, so-called stem-like cancer cells. The purported advantage of these cell is their ability to retain genetics and transcriptional characteristics of the human disease better than standard serum grown human cell lines.^14,18,19^ Examining the effect of novel compounds on these stem-like cells may be more clinically relevant. However, there was no evidence of a smaller (or larger) variance of temozolomide sensitivity compared with other human cell lines. Better understanding of the molecular characteristics of tumour cells as well as experimental conditions in studies using patient-derived cell lines can increase comparability and reproducibility between studies. Initiatives that offer molecularly defined and clinically representative patient-derived cell lines such as The Human Glioblastoma Cell Culture resource^20^ could facilitate more reproducible drug screening.

One of the challenges in summarising temozolomide sensitivities from the included studies was the suboptimal reporting of key experimental conditions. These conditions, such as cell density and carbon dioxide levels, can affect the accuracy of cell viability assay readout.^21^ Guidelines for the conduct of using cell lines for biomedical research^22^ relate to best practice in the standard operating procedure within each laboratory. While some aspects of these guidelines are relevant to the consistency of cancer cell line behaviours, they do not provide a framework for reporting experimental conditions that could affect compound sensitivity. In clinical research, the EQUATOR network has facilitated the development of tailored reporting guidelines specific to different study designs. The earliest guideline was the Consolidated Standards of Reporting Trials (CONSORT) statement^23^, which has been cited for over 8,000 times since its first version published 25 years ago.^24^ The conception of this guideline stemmed from a unanimous opinion from a group of clinical research experts that the quality of reports of randomised controlled trials was in adequate. Our findings from malignant glioma cell line studies indicate similar concerns about the reporting quality in drug discovery studies using cancer cell lines. Variations in experimental design may not be fully captured in a standardised reporting guideline. However, without reproducibility and predictability, data interpretation is unreliable. The issue of reproducibility in pre-clinical animal models that drove the development of ARRIVE guidelines.^12^ What we have in cell line studies should prompt development of guidelines for transparency, consistency and reproducibility.

### Strengths and limitations

This systematic review had a comprehensive search that included all malignant glioma cell lines with their respective drug sensitivity to temozolomide. The vast number of cell lines summarised provided an overview of *in vitro* studies investigating malignant gliomas. Findings of our review also described the methodological practice and reporting associated with the assessment of cell viability to therapeutic agents. All these are important considerations for better design comparable studies in the future.

We drew our results only from malignant glioma cell line models. There are other related experimental models such as drug resistant cell lines, genetically modified cell lines, tumour spheres, organoids and xenotransplant models.^10^ Our findings are likely to be relevant and generalisable to other models because sources of heterogeneity remain uncontrolled and there are more margins of errors associated with other experimental models. The need for consistency and reproducibility apply to all models. There may be other cellular or experimental features that affect cell viability that were not assessed here. These may include *MGMT* promoter methylation, isocitrate dehydrogenase (*IDH*) mutations, telomerase reverse transcriptase (TERT) mutations, 1p/19q co-deletion and alpha-thalassemia mental retardation syndrome (*ATRX*) mutations.^25^ While it would be useful to summarise, there is little consistent evidence to support their role in cell viability and they are very unlikely to be reported in non-patient-derived cell lines. These specific mutations are unlikely to contribute to the observed variability in classical cell lines which should each, in theory, share the same genetic background, although other genetic and epigenetic changes may have been acquired in culture over time.^26^ Studies using patient-derived cell lines often reported patient characteristics, which could partially explain the drug sensitivity variations observed. These cell lines are more likely to be glioma stem cell lines, which are more heterogeneous. The small number of cell lines with comparable experimental conditions would prevent meaningful subgroup analyses to investigate additional the effect of additional characteristics on cell viability.

## Conclusions

Temozolomide sensitivity reported in comparable studies was not consistent between and within individual malignant glioma cell lines. This raises concerns about the reliability and translational value of drug discovery studies that use glioma cell lines. Appraisal and interpretation of the results can only be achievable if studies report key experimental conditions. While there will be variations of opinion on what the optimal design is, a consensus model of a reporting structure is the only rational way to maximise the yield from *in vitro* studies to find novel therapies for our patients.

## Data Availability

Data is available on request

