## Supplementary materials for "Temozolomide sensitivity of malignant glioma cell lines – a systematic review assessing consistencies between *in vitro* studies"

**Supplementary Material**

Supplementary methods – Search strategy

Supplementary references – Citations of 213 included studies in this systematic review

**Search strategy**

Medline

1. (glioblastoma or gbm or (grade adj2 IV adj2 glioma) or (grade adj2 "4" adj2 glioma) or (glioblastoma adj2 multiforme) or glioma*).mp. or exp glioblastoma/ or exp glioma/
2. exp cell line/ or exp tumor cells, cultured/ or (tumo$rstem cell* or cancer* stem cell* or (cell adj2 line) or tumo?r? cell* line$ or patient$derived or (patient adj3 derive*) or (C6 adj3 cell adj2 line) or (GL$261* or 9L$acZ* or F$98* or RG$2* or CNS$1* or C6* or U$251* or U$87* or T$98$G* or LN$229* or SNB$19*) adj3 cell).mp.
3. exp temozolomide/ or (temozolomide or temodar or TMZ).mp
4. exp cell death/ or exp cellular senescence/ or exp cell survival/ or exp drug resistance/ or (survival or killing or autophagy or growth or sensitivity).mp
5. Review/ or editorial/
6. 1-4 not 5

Embase

1. (glioblastoma or gbm or (grade adj2 IV adj2 glioma) or (grade adj2 4 adj2 glioma)
2. exp glioblastoma cell line/ or exp glioblastoma multiforme cell line/ or exp malignant glioma cell line/ or exp glioma cell line/ or exp "C6 cell line (glioma)"/ or (U$251* or U$87* or T$98$G* or LN$229* or SNB$19* or GL$261* or 9L$acZ* or F$98* or RG$2* or CNS$1* or C6*).
3. exp temozolomide/ or (temozolomide or temodar or TMZ).mp
4. exp cell death/ or exp cellular senescence/ or exp cell survival/ or exp drug resistance/ or (survival or killing or autophagy or growth or sensitivity).mp
5. Review/ or editorial/
6. 1-4 not 5

**Supplementary references**

All references are also available at this address: <https://www.zotero.org/groups/4329395/temozolomide_sensitivity_of_malignant_glioma_cell_lines/library>

s1. Aghi M, Rabkin S, Martuza RL. Effect of chemotherapy-induced DNA repair on oncolytic herpes simplex viral replication. *Journal of the National Cancer Institute.* 2006; 98(1):38-50.
s2. Alexiou GA, Gerogianni P, Vartholomatos E, Kyritsis AP. Deferiprone Enhances Temozolomide Cytotoxicity in Glioma Cells. *Cancer investigation.* 2016; 34(10):489-495.
s3. Alonso MM, Gomez-Manzano C, Bekele BN, Yung WKA, Fueyo J. Adenovirus-based strategies overcome temozolomide resistance by silencing the O6-methylguanine-DNA methyltransferase promoter. *Cancer research.* 2007; 67(24):11499-11504.
s4. Annovazzi L, Mellai M, Caldera V, Schiffer D. DNA damage and repair cascade after chemotherapeutic drugs in glioblastoma cell lines. *Anticancer Research.* 2014; 34(10):5811.
s5. Ashizawa T, Miyata H, Iizuka A, et al. Effect of the STAT3 inhibitor STX-0119 on the proliferation of cancer stem-like cells derived from recurrent glioblastoma. *International journal of oncology.* 2013; 43(1):219-227.
s6. Atif F, Patel NR, Yousuf S, Stein DG. The Synergistic Effect of Combination Progesterone and Temozolomide on Human Glioblastoma Cells. *PloS one.* 2015; 10(6):e0131441.
s7. Azambuja JH, Gelsleichter NE, Beckenkamp LR, et al. CD73 Downregulation Decreases In Vitro and In Vivo Glioblastoma Growth. *Molecular neurobiology.* 2019; 56(5):3260-3279.
s8. Balvers RK, Lamfers MLM, Kloezeman JJ, et al. ABT-888 enhances cytotoxic effects of temozolomide independent of MGMT status in serum free cultured glioma cells. *Journal of translational medicine.* 2015; 13(101190741):74.
s9. Balzarotti M, Ciusani E, Calatozzolo C, Croci D, Boiardi A, Salmaggi A. Effect of association of temozolomide with other chemotherapic agents on cell growth inhibition in glioma cell lines. *Oncology research.* 2004; 14(7-8):325-330.
s10. Barazzuol L, Jena R, Burnet NG, et al. Evaluation of poly (ADP-ribose) polymerase inhibitor ABT-888 combined with radiotherapy and temozolomide in glioblastoma. *Radiation oncology (London, England).* 2013; 8(101265111):65.
s11. Barbarisi M, Iaffaioli RV, Armenia E, et al. Novel nanohydrogel of hyaluronic acid loaded with quercetin alone and in combination with temozolomide as new therapeutic tool, CD44 targeted based, of glioblastoma multiforme. *Journal of cellular physiology.* 2018; 233(10):6550-6564.
s12. Beier D, Schriefer B, Brawanski K, et al. Efficacy of clinically relevant temozolomide dosing schemes in glioblastoma cancer stem cell lines. *Journal of neuro-oncology.* 2012; 109(1):45-52.
s13. Berthier S, Arnaud J, Champelovier P, et al. Anticancer properties of sodium selenite in human glioblastoma cell cluster spheroids. *Journal of trace elements in medicine and biology : organ of the Society for Minerals and Trace Elements (GMS).* 2017; 44(b7q, 9508274):161-176.
s14. Berthois Y, Delfino C, Metellus P, et al. Differential expression of miR200a-3p and miR21 in grade II-III and grade IV gliomas: evidence that miR200a-3p is regulated by O6-methylguanine methyltransferase and promotes temozolomide responsiveness. *Cancer biology & therapy.* 2014; 15(7):938-950.
s15. Bobola MS, Kolstoe DD, Blank A, Silber JR. Minimally cytotoxic doses of temozolomide produce radiosensitization in human glioblastoma cells regardless of MGMT expression. *Molecular cancer therapeutics.* 2010; 9(5):1208-1218.
s16. Bobola MS, Tseng SH, Blank A, Berger MS, Silber JR. Role of O6-methylguanine-DNA methyltransferase in resistance of human brain tumor cell lines to the clinically relevant methylating agents temozolomide and streptozotocin. *Clinical cancer research : an official journal of the American Association for Cancer Research.* 1996; 2(4):735-741.
s17. Borges KS, Castro-Gamero AM, Moreno DA, et al. Inhibition of Aurora kinases enhances chemosensitivity to temozolomide and causes radiosensitization in glioblastoma cells. *Journal of cancer research and clinical oncology.* 2012; 138(3):405-414.
s18. Brassesco M, Pezuk JA, Morales AG, et al. Dehydroxymethylepoxyquinomicin efficiently suppresses growth and induces apoptosis in pediatric and adult glioblastoma cell lines. *Pediatric Blood and Cancer.* 2011; 57(5):803-804.
s19. Bruyere C, Abeloos L, Lamoral-Theys D, et al. Temozolomide modifies caveolin-1 expression in experimental malignant gliomas in vitro and in vivo. *Translational Oncology.* 2011; 4(2):92-100.
s20. Cancer M, Drews LF, Bengtsson J, et al. BET and Aurora Kinase A inhibitors synergize against MYCN-positive human glioblastoma cells. *Cell death & disease.* 2019; 10(12):881.
s21. Castro GN, Cayado-Gutierrez N, Zoppino FCM, et al. Effects of temozolomide (TMZ) on the expression and interaction of heat shock proteins (HSPs) and DNA repair proteins in human malignant glioma cells. *Cell stress & chaperones.* 2015; 20(2):253-265.
s22. Castro-Gamero AM, Borges KS, Moreno DA, et al. Tetra-O-methyl nordihydroguaiaretic acid, an inhibitor of Sp1-mediated survivin transcription, induces apoptosis and acts synergistically with chemo-radiotherapy in glioblastoma cells. *Investigational new drugs.* 2013; 31(4):858-870.
s23. Chen C, Han G, Li Y, Yue Z, Wang L, Liu J. FOXO1 associated with sensitivity to chemotherapy drugs and glial-mesenchymal transition in glioma. *Journal of cellular biochemistry.* 2019; 120(1):882-893.
s24. Chen C-M, Syu J-P, Way T-D, et al. BC3EE2,9B, a synthetic carbazole derivative, upregulates autophagy and synergistically sensitizes human GBM8901 glioblastoma cells to temozolomide. *International journal of molecular medicine.* 2015; 36(5):1244-1252.
s25. Chen W, Xu X-K, Li J-L, et al. MALAT1 is a prognostic factor in glioblastoma multiforme and induces chemoresistance to temozolomide through suppressing miR-203 and promoting thymidylate synthase expression. *Oncotarget.* 2017; 8(14):22783-22799.
s26. Chen Y, Gao F, Jiang R, et al. Down-Regulation of AQP4 Expression via p38 MAPK Signaling in Temozolomide-Induced Glioma Cells Growth Inhibition and Invasion Impairment. *Journal of cellular biochemistry.* 2017; 118(12):4905-4913.
s27. Chen Y, Tseng B-J, Tsai Y-H, Tseng S-H. Temozolomide and AZD7762 Induce Synergistic Cytotoxicity Effects on Human Glioma Cells. *Anticancer research.* 2020; 40(9):5141-5149.
s28. Chen Z, Wei X, Shen L, Zhu H, Zheng X. 20(S)-ginsenoside-Rg3 reverses temozolomide resistance and restrains epithelial-mesenchymal transition progression in glioblastoma. *Cancer science.* 2019; 110(1):389-400.
s29. Cheng Q, Ma X, Cao H, et al. Role of miR-223/paired box 6 signaling in temozolomide chemoresistance in glioblastoma multiforme cells. *Molecular medicine reports.* 2017; 15(2):597-604.
s30. Cho H-Y, Wang W, Jhaveri N, et al. NEO212, temozolomide conjugated to perillyl alcohol, is a novel drug for effective treatment of a broad range of temozolomide-resistant gliomas. *Molecular cancer therapeutics.* 2014; 13(8):2004-2017.
s31. Cho H-Y, Wang W, Jhaveri N, et al. Perillyl alcohol for the treatment of temozolomide-resistant gliomas. *Molecular cancer therapeutics.* 2012; 11(11):2462-2472.
s32. Chong DQ, Toh XY, Ho IAW, et al. Combined treatment of Nimotuzumab and rapamycin is effective against temozolomide-resistant human gliomas regardless of the EGFR mutation status. *BMC cancer.* 2015; 15(100967800):255.
s33. Clark PA, Gaal JT, Strebe JK, et al. The effects of tumor treating fields and temozolomide in MGMT expressing and non-expressing patient-derived glioblastoma cells. *Journal of clinical neuroscience : official journal of the Neurosurgical Society of Australasia.* 2017; 36(dpi, 9433352):120-124.
s34. Coelho PLC, Oliveira MN, da Silva AB, et al. The flavonoid apigenin from Croton betulaster Mull inhibits proliferation, induces differentiation and regulates the inflammatory profile of glioma cells. *Anti-cancer drugs.* 2016; 27(10):960-969.
s35. Cousin D, Hummersone MG, Bradshaw TD, et al. Synthesis and growth-inhibitory activities of imidazo?5,1-d]-1,2,3,5-tetrazine-8-carboxamides related to the anti-tumour drug temozolomide, with appended silicon, benzyl and heteromethyl groups at the 3-position. *MedChemComm.* 2018; 9(3):545-553.
s36. Cruickshanks N, Shervington L, Patel R, Munje C, Thakkar D, Shervington A. Can hsp90alpha-targeted siRNA combined with TMZ be a future therapy for glioma? *Cancer investigation.* 2010; 28(6):608-614.
s37. Cui Y, Lin J, Zuo J, et al. AKT2-knockdown suppressed viability with enhanced apoptosis, and attenuated chemoresistance to temozolomide of human glioblastoma cells in vitro and in vivo. *OncoTargets and Therapy.* 2015; 8((Cui, Lin, Zhang, Dong, Hu, Luo, Chen, Lu) Department of Neurosurgery, Changzheng Hospital, Second Military Medical University, Shanghai, China(Zuo) Department of Neurosurgery, First Affiliated Hospital of Soochow University, Suzhou, China):1681-1690.
s38. De Salvo M, Maresca G, D'Agnano I, et al. Temozolomide induced c-Myc-mediated apoptosis via Akt signalling in MGMT expressing glioblastoma cells. *International journal of radiation biology.* 2011; 87(5):518-533.
s39. Ding C, Yi X, Wu X, et al. Exosome-mediated transfer of circRNA CircNFIX enhances temozolomide resistance in glioma. *Cancer letters.* 2020; 479(7600053, cmx):1-12.
s40. Ding L, Wang Q, Shen M, et al. Thermoresponsive nanocomposite gel for local drug delivery to suppress the growth of glioma by inducing autophagy. *Autophagy.* 2017; 13(7):1176-1190.
s41. Doganlar O, Doganlar ZB, Kurtdere AK, Chasan T, Ok ES. Chronic exposure of human glioblastoma tumors to low concentrations of a pesticide mixture induced multidrug resistance against chemotherapy agents. *Ecotoxicology and environmental safety.* 2020; 202(edk, 7805381):110940.
s42. Elhag R, Mazzio EA, Soliman KFA. The effect of silibinin in enhancing toxicity of temozolomide and etoposide in p53 and PTEN-mutated resistant glioma cell lines. *Anticancer research.* 2015; 35(3):1263-1269.
s43. Eugenio M, Campanati L, Muller N, et al. Silver/silver chloride nanoparticles inhibit the proliferation of human glioblastoma cells. *Cytotechnology.* 2018; 70(6):1607-1618.
s44. Fan MD, Zhao XY, Qi JN, et al. TRIM31 enhances chemoresistance in glioblastoma through activation of the PI3K/Akt signaling pathway. *Experimental and Therapeutic Medicine.* 2020; 20(2):802-809.
s45. Feng J, Zhang Y, Ren X, et al. Leucine-rich repeat containing 4 act as an autophagy inhibitor that restores sensitivity of glioblastoma to temozolomide. *Oncogene.* 2020; 39(23):4551-4566.
s46. Finiuk N, Klyuchivska O, Ivasechko I, et al. Proapoptotic effects of novel thiazole derivative on human glioma cells. *Anti-cancer drugs.* 2019; 30(1):27-37.
s47. Fleurence J, Bahri M, Fougeray S, et al. Impairing temozolomide resistance driven by glioma stem-like cells with adjuvant immunotherapy targeting O-acetyl GD2 ganglioside. *International journal of cancer.* 2020; 146(2):424-438.
s48. Forte IM, Indovina P, Iannuzzi CA, et al. Targeted therapy based on p53 reactivation reduces both glioblastoma cell growth and resistance to temozolomide. *International journal of oncology.* 2019; 54(6):2189-2199.
s49. Gaspar N, Marshall L, Perryman L, et al. MGMT-independent temozolomide resistance in pediatric glioblastoma cells associated with a PI3-kinase-mediated HOX/stem cell gene signature. *Cancer research.* 2010; 70(22):9243-9252.
s50. Giacomelli C, Natali L, Trincavelli ML, et al. New insights into the anticancer activity of carnosol: p53 reactivation in the U87MG human glioblastoma cell line. *The international journal of biochemistry & cell biology.* 2016; 74(cdk, 9508482):95-108.
s51. Giussani P, Bassi R, Anelli V, et al. Glucosylceramide synthase protects glioblastoma cells against autophagic and apoptotic death induced by temozolomide and Paclitaxel. *Cancer investigation.* 2012; 30(1):27-37.
s52. Gjika E, Pal-Ghosh S, Kirschner ME, et al. Combination therapy of cold atmospheric plasma (CAP) with temozolomide in the treatment of U87MG glioblastoma cells. *Scientific reports.* 2020; 10(1):16495.
s53. Goellner EM, Grimme B, Brown AR, et al. Overcoming temozolomide resistance in glioblastoma via dual inhibition of NAD+ biosynthesis and base excision repair. *Cancer research.* 2011; 71(6):2308-2317.
s54. Gomez-Manzano C, Lemoine MG, Hu M, et al. Adenovirally-mediated transfer of E2F-1 potentiates chemosensitivity of human glioma cells to temozolomide and BCNU. *International journal of oncology.* 2001; 19(2):359-365.
s55. Grogan PT, Sarkaria JN, Timmermann BN, Cohen MS. Oxidative cytotoxic agent withaferin A resensitizes temozolomide-resistant glioblastomas via MGMT depletion and induces apoptosis through Akt/mTOR pathway inhibitory modulation. *Investigational new drugs.* 2014; 32(4):604-617.
s56. Gu N, Wang X, Di Z, et al. Silencing lncRNA FOXD2-AS1 inhibits proliferation, migration, invasion and drug resistance of drug-resistant glioma cells and promotes their apoptosis via microRNA-98-5p/CPEB4 axis. *Aging.* 2019; 11(22):10266-10283.
s57. Guo X, Luo Z, Xia T, Wu L, Shi Y, Li Y. Identification of miRNA signature associated with BMP2 and chemosensitivity of TMZ in glioblastoma stem-like cells. *Genes and Diseases.* 2020; 7(3):424-439.
s58. Guo Z, Wang H, Wei J, Han L, Li Z. Sequential treatment of phenethyl isothiocyanate increases sensitivity of temozolomide resistant glioblastoma cells by decreasing expression of mgmt via nf-kappab pathway. *American Journal of Translational Research.* 2019; 11(2):696-708.
s59. Han J, Chen Q. MiR-16 modulate temozolomide resistance by regulating BCL-2 in human glioma cells. *International journal of clinical and experimental pathology.* 2015; 8(10):12698-12707.
s60. Hanif F, Perveen K, Malhi SM, Jawed H, Simjee SU. Verapamil potentiates anti-glioblastoma efficacy of temozolomide by modulating apoptotic signaling. *Toxicology in vitro : an international journal published in association with BIBRA.* 2018; 52(8712158, dns):306-313.
s61. Harrabi S, Combs SE, Brons S, Haberer T, Debus J, Weber K-J. Temozolomide in combination with carbon ion or photon irradiation in glioblastoma multiforme cell lines - does scheduling matter? *International journal of radiation biology.* 2013; 89(9):692-697.
s62. He H, Yao M, Zhang W, et al. MEK2 is a prognostic marker and potential chemo-sensitizing target for glioma patients undergoing temozolomide treatment. *Cellular & molecular immunology.* 2016; 13(5):658-668.
s63. Hermisson M, Klumpp A, Wick W, et al. O6-methylguanine DNA methyltransferase and p53 status predict temozolomide sensitivity in human malignant glioma cells. *Journal of neurochemistry.* 2006; 96(3):766-776.
s64. Hiddingh L, Tannous BA, Teng J, et al. EFEMP1 induces gamma-secretase/Notch-mediated temozolomide resistance in glioblastoma. *Oncotarget.* 2014; 5(2):363-374.
s65. Hirose Y, Berger MS, Pieper RO. Abrogation of the Chk1-mediated G2 checkpoint pathway potentiates temozolomide-induced toxicity in a p53-independent manner in human glioblastoma cells. *Cancer Research.* 2001; 61(15):5843-5849.
s66. Howarth A, Simms C, Kerai N, et al. DIVERSet JAG Compounds Inhibit Topoisomerase II and Are Effective Against Adult and Pediatric High-Grade Gliomas. *Translational Oncology.* 2019; 12(10):1375-1385.
s67. Hua L, Huang L, Zhang X, Feng H. Downregulation of hsa_circ_0000936 sensitizes resistant glioma cells to temozolomide by sponging miR-1294. *Journal of Biosciences.* 2020; 45(1):101.
s68. Huang H, Lin H, Zhang X, Li J. Resveratrol reverses temozolomide resistance by downregulation of MGMT in T98G glioblastoma cells by the NF-kappaB-dependent pathway. *Oncology reports.* 2012; 27(6):2050-2056.
s69. Huang X-J, Li C-T, Zhang W-P, Lu Y-B, Fang S-H, Wei E-Q. Dihydroartemisinin potentiates the cytotoxic effect of temozolomide in rat C6 glioma cells. *Pharmacology.* 2008; 82(1):1-9.
s70. Inaba N, Fujioka K, Saito H, et al. Down-regulation of EGFR prolonged cell growth of glioma but did not increase the sensitivity to temozolomide. *Anticancer research.* 2011; 31(10):3253-3257.
s71. Inaba N, Kimura M, Fujioka K, et al. The effect of PTEN on proliferation and drug-, and radiosensitivity in malignant glioma cells. *Anticancer research.* 2011; 31(5):1653-1658.
s72. Inada M, Sato A, Shindo M, et al. Anticancer Non-narcotic Opium Alkaloid Papaverine Suppresses Human Glioblastoma Cell Growth. *Anticancer research.* 2019; 39(12):6743-6750.
s73. Inada M, Shindo M, Kobayashi K, et al. Anticancer effects of a non-narcotic opium alkaloid medicine, papaverine, in human glioblastoma cells. *PloS one.* 2019; 14(5):e0216358.
s74. Jahan N, Lee JM, Shah K, Wakimoto H. Therapeutic targeting of chemoresistant and recurrent glioblastoma stem cells with a proapoptotic variant of oncolytic herpes simplex virus. *International journal of cancer.* 2017; 141(8):1671-1681.
s75. Jayalakshmi J, Vanisree AJ. Naringenin Sensitizes Resistant C6 Glioma Cells with a Repressive Impact on the Migrating Ability. *Annals of Neurosciences.* 2020((Jayalakshmi, Vanisree) Department of Biochemistry, University of Madras, Chennai, Tamil Nadu, India).
s76. Jean-Claude BJ, Mustafa A, Watson AJ, et al. Tetrazepinones are equally cytotoxic to Mer+ and Mer- human tumor cell lines. *The Journal of pharmacology and experimental therapeutics.* 1999; 288(2):484-489.
s77. Jhaveri N, Agasse F, Armstrong D, et al. A novel drug conjugate, NEO212, targeting proneural and mesenchymal subtypes of patient-derived glioma cancer stem cells. *Cancer letters.* 2016; 371(2):240-250.
s78. Johannessen T-CA, Prestegarden L, Grudic A, Hegi ME, Tysnes BB, Bjerkvig R. The DNA repair protein ALKBH2 mediates temozolomide resistance in human glioblastoma cells. *Neuro-oncology.* 2013; 15(3):269-278.
s79. Jun HT, Sun J, Rex K, et al. AMG 102, a fully human anti-hepatocyte growth factor/scatter factor neutralizing antibody, enhances the efficacy of temozolomide or docetaxel in U-87 MG cells and xenografts. *Clinical cancer research : an official journal of the American Association for Cancer Research.* 2007; 13(22 Pt 1):6735-6742.
s80. Kanai R, Rabkin SD, Yip S, et al. Oncolytic virus-mediated manipulation of DNA damage responses: synergy with chemotherapy in killing glioblastoma stem cells. *Journal of the National Cancer Institute.* 2012; 104(1):42-55.
s81. Kanzawa T, Germano IM, Kondo Y, Ito H, Kyo S, Kondo S. Inhibition of telomerase activity in malignant glioma cells correlates with their sensitivity to temozolomide. *British journal of cancer.* 2003; 89(5):922-929.
s82. Karbownik MS, Pietras T, Szemraj J, Kowalczyk E, Nowak JZ. The ability of hyaluronan fragments to reverse the resistance of C6 rat glioma cell line to temozolomide and carmustine. *Wspolczesna Onkologia.* 2014; 18(5):323-328.
s83. Karpel-Massler G, Kast RE, Westhoff M-A, et al. Olanzapine inhibits proliferation, migration and anchorage-independent growth in human glioblastoma cell lines and enhances temozolomide's antiproliferative effect. *Journal of neuro-oncology.* 2015; 122(1):21-33.
s84. Kast RE, Ramiro S, Llado S, Toro S, Covenas R, Munoz M. Antitumor action of temozolomide, ritonavir and aprepitant against human glioma cells. *Journal of neuro-oncology.* 2016; 126(3):425-431.
s85. Kessel S, Cribbes S, Dery O, et al. High-Throughput 3D Tumor Spheroid Screening Method for Cancer Drug Discovery Using Celigo Image Cytometry. *SLAS technology.* 2017; 22(4):454-465.
s86. Khalighfard S, Kalhori MR, Haddad P, Khori V, Alizadeh AM. Enhancement of resistance to chemo-radiation by hsa-miR-1290 expression in glioblastoma cells. *European Journal of Pharmacology.* 2020; 880((Khalighfard, Haddad) Radiation Oncology Research Center, Tehran University of Medical Sciences, Tehran, Iran, Islamic Republic of(Khalighfard, Alizadeh) Breast Disease Research Center, Tehran University of Medical Sciences, Tehran, Iran, Islamic Republic):173144.
s87. Kim SS, Seong S, Lim SH, Kim SY. Biliverdin reductase plays a crucial role in hypoxia-induced chemoresistance in human glioblastoma. *Biochemical and biophysical research communications.* 2013; 440(4):658-663.
s88. Kim S-S, Harford JB, Moghe M, Rait A, Pirollo KF, Chang EH. Targeted nanocomplex carrying siRNA against MALAT1 sensitizes glioblastoma to temozolomide. *Nucleic acids research.* 2018; 46(3):1424-1440.
s89. Kinashi Y, Ikawa T, Takahashi S. The combined effect of neutron irradiation and temozolomide on glioblastoma cell lines with different MGMT and P53 status. *Applied Radiation and Isotopes.* 2020; 163((Kinashi, Ikawa, Takahashi) Institute for Integrated Radiation and Nuclear Science Kyoto University, Kumatori-cho, Sennan-gun, Osaka 590-0494, Japan):109204.
s90. Kobylinska LI, Klyuchivska OY, Grytsyna II, et al. Differential pro-apoptotic effects of synthetic 4-thiazolidinone derivative Les-3288, doxorubicin and temozolomide in human glioma U251 cells. *Croatian medical journal.* 2017; 58(2):150-159.
s91. Kogias E, Osterberg N, Baumer B, et al. Growth-inhibitory and chemosensitizing effects of the glutathione-S-transferase-pi-activated nitric oxide donor PABA/NO in malignant gliomas. *International journal of cancer.* 2012; 130(5):1184-1194.
s92. Krol SK, Bebenek E, Slawinska-Brych A, Dmoszynska-Graniczka M, Boryczka S, Stepulak A. Synthetic Betulin Derivatives Inhibit Growth of Glioma Cells In Vitro. *Anticancer research.* 2020; 40(11):6151-6158.
s93. Kumar DM, Patil V, Ramachandran B, Nila MV, Dharmalingam K, Somasundaram K. Temozolomide-modulated glioma proteome: role of interleukin-1 receptor-associated kinase-4 (IRAK4) in chemosensitivity. *Proteomics.* 2013; 13(14):2113-2124.
s94. Kumar V, Radin D, Leonardi D. Probing the Oncolytic and Chemosensitizing Effects of Dihydrotanshinone in an In Vitro Glioblastoma Model. *Anticancer research.* 2017; 37(11):6025-6030.
s95. Lai IC, Shih P-H, Yao C-J, et al. Elimination of cancer stem-like cells and potentiation of temozolomide sensitivity by Honokiol in glioblastoma multiforme cells. *PloS one.* 2015; 10(3):e0114830.
s96. Lan F, Yang Y, Han J, Wu Q, Yu H, Yue X. Sulforaphane reverses chemo-resistance to temozolomide in glioblastoma cells by NF-kappaB-dependent pathway downregulating MGMT expression. *International journal of oncology.* 2016; 48(2):559-568.
s97. Lan T, Zhao Z, Qu Y, et al. Targeting hyperactivated DNA-PKcs by KU0060648 inhibits glioma progression and enhances temozolomide therapy via suppression of AKT signaling. *Oncotarget.* 2016; 7(34):55555-55571.
s98. Lee C-Y, Lai H-Y, Chiu A, Chan S-H, Hsiao L-P, Lee S-T. The effects of antiepileptic drugs on the growth of glioblastoma cell lines. *Journal of neuro-oncology.* 2016; 127(3):445-453.
s99. Li B, Zhou C, Yi L, Xu L, Xu M. Effect and molecular mechanism of mTOR inhibitor rapamycin on temozolomide-induced autophagic death of U251 glioma cells. *Oncology Letters.* 2018; 15(2):2477-2484.
s100. Li M, Liang RF, Wang X, Mao Q, Liu YH. BKM120 sensitizes C6 glioma cells to temozolomide via suppression of the PI3K/Akt/NF-kappaB/MGMT signaling pathway. *Oncology Letters.* 2017; 14(6):6597-6603.
s101. Li R, Tang D, Zhang J, Wu J, Wang L, Dong J. The temozolomide derivative 2T-P400 inhibits glioma growth via administration route of intravenous injection. *Journal of neuro-oncology.* 2014; 116(1):25-30.
s102. Li R, Xu CQ, Shen JX, et al. 4-Methoxydalbergione is a potent inhibitor of human astroglioma U87 cells in vitro and in vivo. *Acta Pharmacologica Sinica.* 2020((Li, Xu, Shen, Ren, Lin, Huang, Zhong, Luo, Ji, Wu) Brain Function and Disease Laboratory, Shantou University Medical College, Shantou 515041, China(Chen) Guangdong Institute of Microbiology, Guangzhou 510070, China(Li) Yueyang Hospital of Traditional Chi).
s103. Li S, Chen G, Huang J, Li W. MiRNA-27a-3p induces temozolomide resistance in glioma by inhibiting NF1 level. *American Journal of Translational Research.* 2020; 12(8):4749-4756.
s104. Li X-Q, Ouyang Z-G, Zhang S-H, et al. Synergy of enediyne antibiotic lidamycin and temozolomide in suppressing glioma growth with potentiated apoptosis induction. *Journal of neuro-oncology.* 2014; 119(1):91-100.
s105. Li Z-Y, Li Q-Z, Chen L, et al. Histone Deacetylase Inhibitor RGFP109 Overcomes Temozolomide Resistance by Blocking NF-kappaB-Dependent Transcription in Glioblastoma Cell Lines. *Neurochemical research.* 2016; 41(12):3192-3205.
s106. Lin C-J, Lee C-C, Shih Y-L, et al. Resveratrol enhances the therapeutic effect of temozolomide against malignant glioma in vitro and in vivo by inhibiting autophagy. *Free radical biology & medicine.* 2012; 52(2):377-391.
s107. Listik E, Toma L. Glypican-1 in human glioblastoma: Implications in tumorigenesis and chemotherapy. *Oncotarget.* 2020; 11(9):828-845.
s108. Liu Q, Xu X, Zhao M, et al. Berberine induces senescence of human glioblastoma cells by downregulating the EGFR-MEK-ERK signaling pathway. *Molecular cancer therapeutics.* 2015; 14(2):355-363.
s109. Liu Q, Xue Y, Chen Q, et al. PomGnT1 enhances temozolomide resistance by activating epithelial-mesenchymal transition signaling in glioblastoma. *Oncology reports.* 2017; 38(5):2911-2918.
s110. Liu X, Chen J, Li W, Hang C, Dai Y. Inhibition of Casein Kinase II by CX-4945, But Not Yes-associated protein (YAP) by Verteporfin, Enhances the Antitumor Efficacy of Temozolomide in Glioblastoma. *Translational Oncology.* 2020; 13(1):70-78.
s111. Liu X, Wang L, Chen J, et al. Estrogen receptor beta agonist enhances temozolomide sensitivity of glioma cells by inhibiting PI3K/AKT/mTOR pathway. *Molecular medicine reports.* 2015; 11(2):1516-1522.
s112. Liu Y, Xu N, Liu B, et al. Long noncoding RNA RP11-838N2.4 enhances the cytotoxic effects of temozolomide by inhibiting the functions of miR-10a in glioblastoma cell lines. *Oncotarget.* 2016; 7(28):43835-43851.
s113. Liu Y-J, Ma Y-C, Zhang W-J, et al. Combination therapy with micellarized cyclopamine and temozolomide attenuate glioblastoma growth through Gli1 down-regulation. *Oncotarget.* 2017; 8(26):42495-42509.
s114. Luo W, Yan D, Song Z, et al. miR-126-3p sensitizes glioblastoma cells to temozolomide by inactivating Wnt/beta-catenin signaling via targeting SOX2. *Life sciences.* 2019; 226(l62, 0375521):98-106.
s115. Marsoner T, Schmidt OP, Triemer T, Luedtke NW. DNA-Targeted Inhibition of MGMT. *Chembiochem : a European journal of chemical biology.* 2017; 18(10):894-898.
s116. Milani R, Brognara E, Fabbri E, et al. Corilagin induces high levels of apoptosis in the temozolomide-resistant T98G glioma cell line. *Oncology Research.* 2018; 26(9):1307-1315.
s117. Milano V, Piao Y, LaFortune T, de Groot J. Dasatinib-induced autophagy is enhanced in combination with temozolomide in glioma. *Molecular cancer therapeutics.* 2009; 8(2):394-406.
s118. Minaei SE, Khoei S, Khoee S, Vafashoar F, Mahabadi VP. In vitro anti-cancer efficacy of multi-functionalized magnetite nanoparticles combining alternating magnetic hyperthermia in glioblastoma cancer cells. *Materials science & engineering. C, Materials for biological applications.* 2019; 101(101484109):575-587.
s119. Mirabdaly S, Elieh Ali Komi D, Shakiba Y, Moini A, Kiani A. Effects of temozolomide on U87MG glioblastoma cell expression of CXCR4, MMP2, MMP9, VEGF, anti-proliferatory cytotoxic and apoptotic properties. *Molecular biology reports.* 2020; 47(2):1187-1197.
s120. Montaldi AP, Sakamoto-Hojo ET. Methoxyamine sensitizes the resistant glioblastoma T98G cell line to the alkylating agent temozolomide. *Clinical and experimental medicine.* 2013; 13(4):279-288.
s121. Mu Q, Wang L, Yu F, et al. Imp2 regulates GBM progression by activating IGF2/PI3K/Akt pathway. *Cancer biology & therapy.* 2015; 16(4):623-633.
s122. Mullins CS, Schneider B, Stockhammer F, Krohn M, Classen CF, Linnebacher M. Establishment and characterization of primary glioblastoma cell lines from fresh and frozen material: a detailed comparison. *PloS one.* 2013; 8(8):e71070.
s123. Mullins CS, Schubert J, Schneider B, Linnebacher M, Classen CF. Cilengitide response in ultra-low passage glioblastoma cell lines: relation to molecular markers. *Journal of cancer research and clinical oncology.* 2013; 139(8):1425-1431.
s124. Murphy SF, Varghese RT, Lamouille S, et al. Connexin 43 Inhibition Sensitizes Chemoresistant Glioblastoma Cells to Temozolomide. *Cancer research.* 2016; 76(1):139-149.
s125. Musah-Eroje A, Watson S. A novel 3D in vitro model of glioblastoma reveals resistance to temozolomide which was potentiated by hypoxia. *Journal of neuro-oncology.* 2019; 142(2):231-240.
s126. Ni Q, Fan Y, Zhang X, Fan H, Li Y. In vitro and in vivo Study on Glioma Treatment Enhancement by Combining Temozolomide with Calycosin and Formononetin. *Journal of ethnopharmacology.* 2019; 242(7903310, k8t):111699.
s127. Noh H, Zhao Q, Yan J, et al. Cell surface vimentin-targeted monoclonal antibody 86C increases sensitivity to temozolomide in glioma stem cells. *Cancer letters.* 2018; 433(7600053, cmx):176-185.
s128. Pandey V, Ranjan N, Narne P, Babu PP. Roscovitine effectively enhances antitumor activity of temozolomide in vitro and in vivo mediated by increased autophagy and Caspase-3 dependent apoptosis. *Scientific reports.* 2019; 9(1):5012.
s129. Park M, Song C, Yoon H, Choi K-H. Double Blockade of Glioma Cell Proliferation and Migration by Temozolomide Conjugated with NPPB, a Chloride Channel Blocker. *ACS chemical neuroscience.* 2016; 7(3):275-285.
s130. Pedeboscq S, L'Azou B, Passagne I, et al. Cytotoxic and apoptotic effects of bortezomib and gefitinib compared to alkylating agents on human glioblastoma cells. *Journal of experimental therapeutics & oncology.* 2008; 7(2):99-111.
s131. Polewski MD, Reveron-Thornton RF, Cherryholmes GA, Marinov GK, Cassady K, Aboody KS. Increased Expression of System xc- in Glioblastoma Confers an Altered Metabolic State and Temozolomide Resistance. *Molecular cancer research : MCR.* 2016; 14(12):1229-1242.
s132. Poon M-W, Zhuang JTF, Wong STS, Sun S, Zhang X-Q, Leung GKK. Co-expression of Cytoskeletal Protein Adducin 3 and CD133 in Neurospheres and a Temozolomide-resistant Subclone of Glioblastoma. *Anticancer research.* 2015; 35(12):6487-6495.
s133. Potschke R, Gielen G, Pietsch T, et al. Musashi1 enhances chemotherapy resistance of pediatric glioblastoma cells in vitro. *Pediatric Research.* 2020; 87(4):669-676.
s134. Qian X, Ren Y, Shi Z, et al. Sequence-dependent synergistic inhibition of human glioma cell lines by combined temozolomide and miR-21 inhibitor gene therapy. *Molecular pharmaceutics.* 2012; 9(9):2636-2645.
s135. Qiu Z-K, Shen D, Chen Y-S, et al. Valproic acid downregulates the expression of MGMT resistance in glioma stem-like cells. *Chinese journal of cancer.* 2014; 33(2):115-122.
s136. Rainov NG, Fels C, Droege JW, Schafer C, Kramm CM, Chou TC. Temozolomide enhances herpes simplex virus thymidine kinase/ganciclovir therapy of malignant glioma. *Cancer gene therapy.* 2001; 8(9):662-668.
s137. Ramachandran C, Lollett IV, Escalon E, Quirin K-W, Melnick SJ. Anticancer potential and mechanism of action of mango ginger (Curcuma amada Roxb.) supercritical CO2 extract in human glioblastoma cells. *Journal of evidence-based complementary & alternative medicine.* 2015; 20(2):109-119.
s138. Ramachandran C, Nair SM, Escalon E, Melnick SJ. Potentiation of etoposide and temozolomide cytotoxicity by curcumin and turmeric force TM in brain tumor cell lines. *Journal of complementary & integrative medicine.* 2012; 9(101313855):Article-20.
s139. Ramalho MJ, Sevin E, Gosselet F, et al. Receptor-mediated PLGA nanoparticles for glioblastoma multiforme treatment. *International journal of pharmaceutics.* 2018; 545(1-2):84-92.
s140. Ramirez YP, Mladek AC, Phillips RM, et al. Evaluation of novel imidazotetrazine analogues designed to overcome temozolomide resistance and glioblastoma regrowth. *Molecular cancer therapeutics.* 2015; 14(1):111-119.
s141. Ren H, Tan X, Dong Y, et al. Differential effect of imatinib and synergism of combination treatment with chemotherapeutic agents in malignant glioma cells. *Basic & clinical pharmacology & toxicology.* 2009; 104(3):241-252.
s142. Renziehausen A, Tsiailanis AD, Perryman R, et al. Encapsulation of Temozolomide in a Calixarene Nanocapsule Improves Its Stability and Enhances Its Therapeutic Efficacy against Glioblastoma. *Molecular cancer therapeutics.* 2019; 18(9):1497-1505.
s143. Rotondo R, Oliva MA, Staffieri S, Castaldo S, Giangaspero F, Arcella A. Implication of Lactucopicrin in Autophagy, Cell Cycle Arrest and Oxidative Stress to Inhibit U87Mg Glioblastoma Cell Growth. *Molecules (Basel, Switzerland).* 2020; 25(24).
s144. Rubino S, Bach MD, Schober AL, Lambert IH, Mongin AA. Downregulation of leucine-rich repeat-containing 8A limits proliferation and increases sensitivity of glioblastoma to temozolomide and carmustine. *Frontiers in Oncology.* 2018; 8(MAY):142.
s145. Ryu CH, Yoon WS, Park KY, et al. Valproic acid downregulates the expression of MGMT and sensitizes temozolomide-resistant glioma cells. *Journal of biomedicine & biotechnology.* 2012; 2012(101135740):987495.
s146. Saito R, Bringas JR, Panner A, et al. Convection-enhanced delivery of tumor necrosis factor-related apoptosis-inducing ligand with systemic administration of temozolomide prolongs survival in an intracranial glioblastoma xenograft model. *Cancer research.* 2004; 64(19):6858-6862.
s147. Sankar A, Thomas DG, Darling JL. Sensitivity of short-term cultures derived from human malignant glioma to the anti-cancer drug temozolomide. *Anti-cancer drugs.* 1999; 10(2):179-185.
s148. Senbabaoglu F, Aksu AC, Cingoz A, et al. Drug Repositioning Screen on a New Primary Cell Line Identifies Potent Therapeutics for Glioblastoma. *Frontiers in Neuroscience.* 2020; 14((Senbabaoglu, Aksu, Cingoz, Seker-Polat, Bagci-Onder) Brain Cancer Research and Therapy Laboratory, Koc University School of Medicine, Istanbul, Turkey(Senbabaoglu, Aksu, Cingoz, Seker-Polat, Solaroglu, Bagci-Onder) Koc University Research Center for Tran):578316.
s149. Shangguan W, Lv X, Tian N. FoxD2-AS1 is a prognostic factor in glioma and promotes temozolomide resistance in a O6-methylguanine-DNA methyltransferase-dependent manner. *Korean Journal of Physiology and Pharmacology.* 2019; 23(6):475-482.
s150. Shao C-j, Wu M-w, Chen F-r, Li C, Xia Y-f, Chen Z-p. Histone deacetylase inhibitor, 2-propylpentanoic acid, increases the chemosensitivity and radiosensitivity of human glioma cell lines in vitro. *Chinese medical journal.* 2012; 125(24):4338-4343.
s151. Shervington A, Patel R. Silencing DNA methyltransferase (DNMT) enhances glioma chemosensitivity. *Oligonucleotides.* 2008; 18(4):365-374.
s152. Shi L, Li H, Zhan Y. All-trans retinoic acid enhances temozolomide-induced autophagy in human glioma cells U251 via targeting Keap1/Nrf2/ARE signaling pathway. *Oncology Letters.* 2017; 14(3):2709-2714.
s153. Silva AO, Dalsin E, Onzi GR, Filippi-Chiela EC, Lenz G. The regrowth kinetic of the surviving population is independent of acute and chronic responses to temozolomide in glioblastoma cell lines. *Experimental cell research.* 2016; 348(2):177-183.
s154. Silva VAO, Rosa MN, Miranda-Goncalves V, et al. Euphol, a tetracyclic triterpene, from Euphorbia tirucalli induces autophagy and sensitizes temozolomide cytotoxicity on glioblastoma cells. *Investigational new drugs.* 2019; 37(2):223-237.
s155. Skarkova V, Krupova M, Vitovcova B, et al. The Evaluation of Glioblastoma Cell Dissociation and Its Influence on Its Behavior. *International journal of molecular sciences.* 2019; 20(18).
s156. Song Y, Hu Z, Long H, et al. A complex mechanism for HDGF-mediated cell growth, migration, invasion, and TMZ chemosensitivity in glioma. *Journal of neuro-oncology.* 2014; 119(2):285-295.
s157. Sooman L, Ekman S, Andersson C, et al. Synergistic interactions between camptothecin and EGFR or RAC1 inhibitors and between imatinib and Notch signaling or RAC1 inhibitors in glioblastoma cell lines. *Cancer chemotherapy and pharmacology.* 2013; 72(2):329-340.
s158. Sravya P, Nimbalkar VP, Kanuri NN, et al. Low mitochondrial DNA copy number is associated with poor prognosis and treatment resistance in glioblastoma. *Mitochondrion.* 2020; 55((Sravya) Department of Clinical Neurosciences, National Institute of Mental Health and Neurosciences, Bengaluru, India(Nimbalkar, Kanuri, Sugur, Rao, Santosh) Department of Neuropathology, National Institute of Mental Health and Neurosciences, Bengaluru):154-163.
s159. Stepanenko AA, Dmitrenko VV. Pitfalls of the MTT assay: Direct and off-target effects of inhibitors can result in over/underestimation of cell viability. *Gene.* 2015; 574(2):193-203.
s160. Sun S, Lee D, Lee NP, et al. Hyperoxia resensitizes chemoresistant human glioblastoma cells to temozolomide. *Journal of neuro-oncology.* 2012; 109(3):467-475.
s161. Suzuki Y, Fujioka K, Ikeda K, Murayama Y, Manome Y. Temozolomide does not influence the transcription or activity of matrix metalloproteinases 9 and 2 in glioma cell lines. *Journal of clinical neuroscience : official journal of the Neurosurgical Society of Australasia.* 2017; 41(dpi, 9433352):144-149.
s162. Svec RL, Furiassi L, Skibinski CG, Fan TM, Riggins GJ, Hergenrother PJ. Tunable Stability of Imidazotetrazines Leads to a Potent Compound for Glioblastoma. *ACS chemical biology.* 2018; 13(11):3206-3216.
s163. Szeliga M, Karpinska M, Rola R, Niewiadomy A. Design, synthesis and biological evaluation of novel 1,3,4-thiadiazole derivatives as anti-glioblastoma agents targeting the AKT pathway. *Bioorganic Chemistry.* 2020; 105((Szeliga) Department of Neurotoxicology, Mossakowski Medical Research Centre, Polish Academy of Sciences, 5 Pawinskiego Str, Warsaw 02-106, Poland(Karpinska, Niewiadomy) Lukasiewicz Research Network - Institute of Industrial Organic Chemistry, 6 Annopol S):104362.
s164. Taspinar M, Ilgaz S, Ozdemir M, et al. Effect of lomeguatrib-temozolomide combination on MGMT promoter methylation and expression in primary glioblastoma tumor cells. *Tumour biology : the journal of the International Society for Oncodevelopmental Biology and Medicine.* 2013; 34(3):1935-1947.
s165. Tentori L, Ricci-Vitiani L, Muzi A, et al. Pharmacological inhibition of poly(ADP-ribose) polymerase-1 modulates resistance of human glioblastoma stem cells to temozolomide. *BMC cancer.* 2014; 14(100967800):151.
s166. Tian T, Li A, Lu H, Luo R, Zhang M, Li Z. TAZ promotes temozolomide resistance by upregulating MCL-1 in human glioma cells. *Biochemical and biophysical research communications.* 2015; 463(4):638-643.
s167. Towner RA, Smith N, Saunders D, et al. OKN-007 Increases temozolomide (TMZ) Sensitivity and Suppresses TMZ-Resistant Glioblastoma (GBM) Tumor Growth. *Translational Oncology.* 2019; 12(2):320-335.
s168. Tsai, Tsai N-M, Chang K-F, Wang J-C. Juniperus Communis Extract Exerts Antitumor Effects in Human Glioblastomas Through Blood-Brain Barrier. *Cellular physiology and biochemistry : international journal of experimental cellular physiology, biochemistry, and pharmacology.* 2018; 49(6):2443-2462.
s169. Vaglini F, Pardini C, Di Desidero T, et al. Melanocortin Receptor-4 and Glioblastoma Cells: Effects of the Selective Antagonist ML00253764 Alone and in Combination with Temozolomide In Vitro and In Vivo. *Molecular neurobiology.* 2018; 55(6):4984-4997.
s170. Vengoji R, Macha MA, Nimmakayala RK, et al. Afatinib and Temozolomide combination inhibits tumorigenesis by targeting EGFRvIII-cMet signaling in glioblastoma cells. *Journal of experimental & clinical cancer research : CR.* 2019; 38(1):266.
s171. Villalva C, Cortes U, Wager M, et al. O6-Methylguanine-methyltransferase (MGMT) promoter methylation status in glioma stem-like cells is correlated to temozolomide sensitivity under differentiation-promoting conditions. *International journal of molecular sciences.* 2012; 13(6):6983-6994.
s172. Viswanathan A, Sebastianelli G, Brown K, et al. In vitro anti-glioblastoma activity of L-valine derived boroxazolidones. *European journal of pharmacology.* 2019; 854(en6, 1254354):194-200.
s173. Wang H, Cai S, Bailey BJ, et al. Combination therapy in a xenograft model of glioblastoma: Enhancement of the antitumor activity of temozolomide by an MDM2 antagonist. *Journal of Neurosurgery.* 2017; 126(2):446-459.
s174. Wang J, Sai K, Chen F-r, Chen Z-p. Oroxylin A increases the sensitivity of temozolomide to temozolomide by targeting MEK1. *Cancer chemotherapy and pharmacology.* 2013; 72(1):147-158.
s175. Wang L, Tang S, Yu Y, et al. Intranasal Delivery of Temozolomide-Conjugated Gold Nanoparticles Functionalized with Anti-EphA3 for Glioblastoma Targeting. *Molecular Pharmaceutics.* 2021((Wang, Tang, Yu, Lv, Wang, Yan, Li, Sun, Li) School of Pharmacy, Key Laboratory of Molecular Pharmacology and Drug Evaluation (Yantai University), Ministry of Education, Collab. Innov. Center of Advanced Drug Delivery System and Biotech Drugs in Universit).
s176. Wang Q, Qian J, Wang J, et al. Knockdown of RLIP76 expression by RNA interference inhibits invasion, induces cell cycle arrest, and increases chemosensitivity to the anticancer drug temozolomide in glioma cells. *Journal of neuro-oncology.* 2013; 112(1):73-82.
s177. Wang W, Han S, Gao W, Feng Y, Li K, Wu D. Long Noncoding RNA KCNQ1OT1 Confers Gliomas Resistance to Temozolomide and Enhances Cell Growth by Retrieving PIM1 From miR-761. *Cellular and Molecular Neurobiology.* 2020((Wang, Han, Gao, Feng, Li) Department of Neurosurgery, The First Affiliated Hospital of China Medical University, Shenyang, Liaoning 110001, China(Wu) Department of Tumor Biotherapy and Cancer Research, The First Affiliated Hospital of China Medical Unive).
s178. Wang X, Chen JX, Liu JP, You C, Liu YH, Mao Q. Gain of function of mutant TP53 in glioblastoma: Prognosis and response to temozolomide. *Annals of Surgical Oncology.* 2014; 21(4):1337-1344.
s179. Wang X, Chen J-x, Liu Y-h, You C, Mao Q. Mutant TP53 enhances the resistance of glioblastoma cells to temozolomide by up-regulating O(6)-methylguanine DNA-methyltransferase. *Neurological sciences : official journal of the Italian Neurological Society and of the Italian Society of Clinical Neurophysiology.* 2013; 34(8):1421-1428.
s180. Wang X, Jia L, Jin X, et al. NF-kappaB inhibitor reverses temozolomide resistance in human glioma TR/U251 cells. *Oncology Letters.* 2015; 9(6):2586-2590.
s181. Wei M, Ma R, Huang S, et al. Oroxylin A increases the sensitivity of temozolomide on glioma cells by hypoxia-inducible factor 1alpha/hedgehog pathway under hypoxia. *Journal of Cellular Physiology.* 2019; 234(10):17392-17404.
s182. Wei YT, Yang QY, Fan SB, Liang W, Hu DX, Shuai P. Sinomenine inhibits the growth of glioma cells through STAT3 signal pathway. *Journal of Applied Biomedicine.* 2018; 16(1):22-28.
s183. Weiss T, Schneider H, Silginer M, et al. NKG2D-Dependent Antitumor Effects of Chemotherapy and Radiotherapy against Glioblastoma. *Clinical cancer research : an official journal of the American Association for Cancer Research.* 2018; 24(4):882-895.
s184. Wen X, Li S, Guo M, et al. miR-181a-5p inhibits the proliferation and invasion of drug-resistant glioblastoma cells by targeting F-box protein 11 expression. *Oncology Letters.* 2020; 20(5):12098.
s185. Wu Y, Dong L, Bao S, Wang M, Yun Y, Zhu R. FK228 augmented temozolomide sensitivity in human glioma cells by blocking PI3K/AKT/mTOR signal pathways. *Biomedicine & pharmacotherapy = Biomedecine & pharmacotherapie.* 2016; 84(a59, 8213295):462-469.
s186. Wu Y, Yao Y, Yun Y, Wang M, Zhu R. MicroRNA-302c enhances the chemosensitivity of human glioma cells to temozolomide by suppressing P-gp expression. *Bioscience reports.* 2019; 39(9).
s187. Xiao S, Yang Z, Qiu X, et al. miR-29c contribute to glioma cells temozolomide sensitivity by targeting O6-methylguanine-DNA methyltransferases indirectely. *Oncotarget.* 2016; 7(31):50229-50238.
s188. Xiong Y, Chen R, Wang L, et al. Downregulation of miR-186 promotes the proliferation and drug resistance of glioblastoma cells by targeting Twist1. *Molecular medicine reports.* 2019; 19(6):5301-5308.
s189. Xu P, Zhang G, Hou S, Sha L-G. MAPK8 mediates resistance to temozolomide and apoptosis of glioblastoma cells through MAPK signaling pathway. *Biomedicine & pharmacotherapy = Biomedecine & pharmacotherapie.* 2018; 106(a59, 8213295):1419-1427.
s190. Xu Y, Shen M, Li Y, et al. The synergic antitumor effects of paclitaxel and temozolomide co-loaded in mPEG-PLGA nanoparticles on glioblastoma cells. *Oncotarget.* 2016; 7(15):20890-20901.
s191. Yamada T, Tsuji S, Nakamura S, et al. Riluzole enhances the antitumor effects of temozolomide via suppression of MGMT expression in glioblastoma. *Journal of neurosurgery.* 2020((Yamada, Tsuji, Nakamura, Shimazawa, Hara) 1Molecular Pharmacology, Department of Biofunctional Evaluation, Gifu Pharmaceutical University; and(Yamada, Egashira, Nakayama, Yano, Iwama) Department of Neurosurgery, Gifu University Graduate School of Medicin):1-10.
s192. Yan Y, Xu Z, Chen X, et al. Novel Function of lncRNA ADAMTS9-AS2 in Promoting Temozolomide Resistance in Glioblastoma via Upregulating the FUS/MDM2 Ubiquitination Axis. *Frontiers in Cell and Developmental Biology.* 2019; 7((Yan, Chen, Wang, Zeng, Qian, Wei, Gong) Department of Pharmacy, Xiangya Hospital, Central South University, Changsha, China(Yan, Chen, Wang, Zeng, Qian, Wei, Gong) National Clinical Research Center for Geriatric Disorders, Xiangya Hospital, Central South):217.
s193. Yang Q, Deng L, Li J, Miao P, Liu W, Huang Q. Nr5a2 promotes cell growth and resistance to temozolomide through regulating notch signal pathway in glioma. *OncoTargets and Therapy.* 2020; 13((Yang, Li, Miao, Liu, Huang) Department of Neurosurgery, The First People's Hospital of Shangqiu in Henan Province, Shangqiu Clinical College, Xuzhou Medical University, Shangqiu 476100, China(Deng) Department of Neonatology, The First People's Hospital o):10231-10244.
s194. Yang SH, Li S, Lu G, et al. Metformin treatment reduces temozolomide resistance of glioblastoma cells. *Oncotarget.* 2016; 7(48):78787-78803.
s195. Yin H, Zhou Y, Wen C, et al. Curcumin sensitizes glioblastoma to temozolomide by simultaneously generating ROS and disrupting AKT/mTOR signaling. *Oncology reports.* 2014; 32(4):1610-1616.
s196. Yoshino A, Ogino A, Yachi K, et al. Gene expression profiling predicts response to temozolomide in malignant gliomas. *International journal of oncology.* 2010; 36(6):1367-1377.
s197. Yu G, Wu F, Wang E. KLF8 Promotes Temozolomide Resistance in Glioma Cells via beta-Catenin Activation. *Cellular physiology and biochemistry : international journal of experimental cellular physiology, biochemistry, and pharmacology.* 2016; 38(4):1596-1604.
s198. Yu Q, Liu L, Wang P, Yao Y, Xue Y, Liu Y. EMAP-II sensitize U87MG and glioma stem-like cells to temozolomide via induction of autophagy-mediated cell death and G2/M arrest. *Cell cycle (Georgetown, Tex.).* 2017; 16(11):1085-1092.
s199. Yun EJ, Kim S, Hsieh JT, Baek ST. Wnt/beta-catenin signaling pathway induces autophagy-mediated temozolomide-resistance in human glioblastoma. *Cell Death and Disease.* 2020; 11(9):771.
s200. Zhang D, Dai D, Zhou M, et al. Inhibition of Cyclin D1 Expression in Human Glioblastoma Cells is Associated with Increased Temozolomide Chemosensitivity. *Cellular physiology and biochemistry : international journal of experimental cellular physiology, biochemistry, and pharmacology.* 2018; 51(6):2496-2508.
s201. Zhang J, Stevens MFG, Hummersone M, Madhusudan S, Laughton CA, Bradshaw TD. Certain imidazotetrazines escape O6-methylguanine-DNA methyltransferase and mismatch repair. *Oncology.* 2011; 80(3-4):195-207.
s202. Zhang J, Stevens MFG, Laughton CA, Madhusudan S, Bradshaw TD. Acquired resistance to temozolomide in glioma cell lines: molecular mechanisms and potential translational applications. *Oncology.* 2010; 78(2):103-114.
s203. Zhang J, Wang Z, Ye Y, et al. Glioma cells with Tim-3 expression induce resistance to chemotherapy drug. *International Journal of Clinical and Experimental Medicine.* 2018; 11(12):13442-13447.
s204. Zhang L, Yan Y, Jiang Y, et al. Knockdown of SALL4 expression using RNA interference induces cell cycle arrest, enhances early apoptosis, inhibits invasion and increases chemosensitivity to temozolomide in U251 glioma cells. *Oncology Letters.* 2017; 14(4):4263-4269.
s205. Zhang P, Chen X-B, Ding B-Q, Liu H-L, He T. Down-regulation of ABCE1 inhibits temozolomide resistance in glioma through the PI3K/Akt/NF-kappaB signaling pathway. *Bioscience reports.* 2018; 38(6).
s206. Zhang P, Tang M, Huang Q, et al. Combination of 3-methyladenine therapy and Asn-Gly-Arg (NGR)-modified mesoporous silica nanoparticles loaded with temozolomide for glioma therapy in vitro. *Biochemical and Biophysical Research Communications.* 2019; 509(2):549-556.
s207. Zhang X, Yu J, Zhao C, et al. MiR-181b-5p modulates chemosensitivity of glioma cells to temozolomide by targeting Bcl-2. *Biomedicine & pharmacotherapy = Biomedecine & pharmacotherapie.* 2019; 109(a59, 8213295):2192-2202.
s208. Zhang Y, Wang S-X, Ma J-W, et al. EGCG inhibits properties of glioma stem-like cells and synergizes with temozolomide through downregulation of P-glycoprotein inhibition. *Journal of neuro-oncology.* 2015; 121(1):41-52.
s209. Zhao M, Bozzato E, Joudiou N, et al. Codelivery of paclitaxel and temozolomide through a photopolymerizable hydrogel prevents glioblastoma recurrence after surgical resection. *Journal of controlled release : official journal of the Controlled Release Society.* 2019; 309(c46, 8607908):72-81.
s210. Zhou B, Bu G, Zhou Y, Zhao Y, Li W, Li M. Knockdown of CDC2 expression inhibits proliferation, enhances apoptosis, and increases chemosensitivity to temozolomide in glioblastoma cells. *Medical oncology (Northwood, London, England).* 2015; 32(1):378.
s211. Zhu D, Tu M, Zeng B, et al. Up-regulation of miR-497 confers resistance to temozolomide in human glioma cells by targeting mTOR/Bcl-2. *Cancer medicine.* 2017; 6(2):452-462.
s212. Zhu Z, Zhang P, Li N, et al. Lovastatin Enhances Cytotoxicity of Temozolomide via Impairing Autophagic Flux in Glioblastoma Cells. *BioMed research international.* 2019; 2019(101600173):2710693.
